# Barriers and Facilitators in Implementing Online Educational Interventions for Physicians: Systematic Review of Reviews

**DOI:** 10.1101/2024.11.18.24317472

**Authors:** Akbota Tolegenova, Akbota Kanderzhanova, Faye Foster, Paolo Colet, Valentina Stolyarova, Antonio Sarria-Santamera

**Affiliations:** Department of Medicine, Nazarbayev University School of Medicine, 5/1 Kerey and Zhanibek Khans St, Astana 020000, Kazakhstan

**Keywords:** Nurses, Education, Autism

## Abstract

The implementation of online educational interventions for physicians presents both barriers and facilitators that shape the integration of digital learning into medical education frameworks. This systematic review of reviews aims to synthesize evidence on factors influencing the successful implementation of online educational interventions for physicians. We conducted searches across databases including, PubMed/Medline, Cochrane, Web of Science, Embase, PsycInfo, CINAHL spanning the years 2018 to 2023. Inclusion criteria comprised systematic reviews focusing on the implementation of e-learning. Data synthesis followed meta-ethnography principles, with categorization guided by the Consolidated Framework for Implementation Research (CFIR). Twenty-one relevant reviews were analyzed, highlighting factors within the CFIR domains: Innovation, Outer Setting, Inner Setting, Individuals, and Implementation Process. Facilitators included evidence-based practices, adaptability, leadership engagement, and resource availability, while barriers encompassed limited funding, regulatory hurdles, technological constraints, and resistance to change. The results emphasize how crucial it is for healthcare institutions, policymakers, educators, and technology providers to work together in order to effectively incorporate online learning into medical education. In conclusion, the study highlights the importance of tailored implementation strategies and policy recommendations, utilizing evidence-based practices and addressing various factors, to enhance online medical education’s effectiveness and ensure its integration into healthcare systems for improved patient outcomes.

---

Online learning has revolutionized education globally, providing students with accessible and flexible learning opportunities that extend beyond academics to encompass extracurricular activities (Hiltz & Turoff, 2005). From interactive content and global collaboration to adaptive learning and teacher professional development, online platforms have become indispensable tools for modern education, shaping a dynamic and inclusive learning environment (Haleem et al., 2022).

## Online Education

Defined by Singh and Thurman (2019) as the delivery of educational content through the internet, online education, allows students to learn independently of their physical or virtual location. Instructors create online teaching modules to enhance interactivity in both synchronous and asynchronous settings. It encompasses the utilization of technological advancements to guide, create, and convey educational content while enabling interactive communication between students and instructors (Thanji & Vasantha, 2016).

Moreover, it was found that online learning offers the advantages of flexibility, accessibility, and personalized learning experiences, empowering students to engage with educational content at their own pace and convenience (Mukhtar et al., 2020). According to the National Center for Education Statistics (2022), in the fall of 2021, approximately 60% of college students, totaling 11,205,320 individuals nationwide, enrolled in at least one online course across the USA. The substantial increase in online enrollment is indicative of a rising pattern in the changing realm of education, particularly within the healthcare profession.

## Physicians and Online Education

This surge in online education is crucial given the projections by the World Health Organization (WHO) (2016), which anticipates a potential deficit of 10 million health workers by 2030. This shortage poses significant challenges in delivering effective healthcare services, particularly in developing countries where limited faculty and institutional resources compound existing issues in healthcare quality (Berendes et al., 2011).

Adding to the difficulty, medical staff often face deficient knowledge and skills, exacerbated by a widening gap between the rapid advances and innovations in healthcare and their effective dissemination to frontline professionals, especially physicians working in primary health centers (Pakenham-Walsh & Bukachi, 2009). This knowledge and skill disparity creates a significant obstacle in maintaining a high standard of healthcare provision (Mosadeghrad, 2014).

To address this critical issue, various healthcare fraternities have launched comprehensive training programs, prominently featuring continuing professional development and continuing medical education initiatives (Davis et al., 2008). These programs aim not only to link the existing knowledge and skill gap but also to allow healthcare professionals, particularly physicians, with a latest advancements in approaches and best practices in their respective fields (Ahuja, 2019). Through these initiatives, professionals are better equipped to meet the evolving demands of the healthcare landscape, contributing to an overall improvement in the quality of healthcare services (Mosadeghrad, 2014).

One way to help physicians meet evolving demands is through online educational trainings. Those web-based educational interventions offer a cost-effective, easily implementable, and accessible approach for healthcare professionals, providing a platform for effective learning and skill enhancement (Fredericks et al., 2014). Numerous studies have demonstrated the efficacy of internet-based educational interventions across various medical topics (Martinić et al., 2022; Laine et al., 2019). Moreover, a thorough evaluation of 26 studies on online educational interventions for physicians revealed that such interventions consistently result in a significant improvement in the participants’ knowledge levels. This finding implies a positive relationship between engaging in online health education and an increase of knowledge among participants (Claflin et al., 2021).

## Factors Influencing Implementation

Among this paradigm shift, examining the factors influencing the successful implementation of online educational interventions for physicians becomes crucial. While the advantages of online education are apparent, challenges and barriers may hinder the seamless integration of these interventions into the established medical education framework (O’Doherty et al., 2018). Technological constraints, such as limited access to reliable internet connectivity or outdated hardware, can hinder the effectiveness of online learning experiences, especially in low-middle income countries (Adedoyın & Soykan, 2020). Institutional resistance and lack of support can also hinder online intervention implementation (Bury et al., 2006). Additionally, understanding physician preferences and addressing concerns about the efficacy of online learning methodologies are also found to be crucial for fostering acceptance and engagement (Ismail et al., 2021).

## Study Rationale and Aim

With numerous reviews covering different factors of online educational interventions, it can be challenging for researchers, healthcare managers, or policymakers to find and apply relevant evidence that fits their specific needs (Ross et al., 2016). Existing reviews have explored various aspects of online medical education, including its impact on knowledge acquisition, attitudes, skills, and patient outcomes (Lawn et al., 2017). However, synthesizing this information is crucial to provide a nuanced understanding of the barriers and facilitators that shape the implementation factors of online educational interventions for physicians. These insights are vital for developing effective strategies to successfully integrate online intervention into healthcare, contributing valuable perspectives to the ongoing discussions about the future of physicians’ knowledge in the digital age. This study is a systematic review of reviews. Its primary aim is to identify and synthesize evidence from existing reviews on barriers and facilitators in implementing online educational interventions for physicians.

## Methods

This study is a systematic review of reviews, which is a comprehensive research method that involves synthesizing and analyzing the findings of multiple systematic reviews on a particular topic or research question and defined by Smith et al. (2011). According to his methodology, this study design aggregates and assesses the evidence presented in various systematic reviews and allows researchers to gain a broader perspective, considering a range of interventions, outcomes, populations, or settings. This systematic review has been registered with PROSPERO under the registration number CRD42024589492.

### Inclusion and Exclusion Criteria

The PICOS (Population, Intervention, Control, Outcomes, and Study Type) strategy was systematically employed to construct the eligibility criteria for the inclusion and exclusion of studies in this review (Amir-Behghadami & Janati, 2020).

Population: The target population comprised healthcare professionals across various disciplines, including but not limited to physicians, nurses, pharmacists, and allied health professionals. Studies involving participants within the healthcare profession were included.

Intervention: The internet-based educational interventions designed for healthcare professionals encompassed various e-health initiatives, including online courses, telemedicine training, web-based modules, and digital resources to enhance professional knowledge, skills, awareness, services, and communication.

Control: This study was not restricted to comparator studies.

Outcome: The outcomes of interest included qualitative data on factors influencing the implementation of internet-based educational interventions, both facilitating and hindering aspects. Additionally, the focus was on strategies reported in the literature to promote the successful implementation of e-health initiatives among healthcare professions.

Study type: The study type included systematic reviews, meta-analyses, qualitative meta-syntheses, thematic synthesis and meta-ethnographies written in English. These study designs were chosen to ensure a comprehensive overview of the existing literature and to facilitate the synthesis of evidence from multiple sources. Primary research studies, secondary analyses, commentaries, and editorials were excluded to maintain the focus on aggregated findings.

### Search Strategy

To ensure an exhaustive search, a comprehensive strategy was developed. Cochrane Library, Web of Science, PubMed/Medline, EMBASE, PSCHINFO databases were systematically searched using MESH and Emtree. CINAHL database was also utilized to search for grey literature. Boolean operators (AND, OR) were employed to combine relevant terms to enhance search precision. Search strings were as following:

1. (‘Internet-based education’ OR ‘Online education’ OR ‘E-learning’ OR ‘Web-based training’ OR ‘Electronic health education’ OR ‘Digital learning’ OR ‘Internet interventions’ OR ‘Telemedicine education’) AND (‘Healthcare professionals’ OR ‘Medical professionals’ OR ‘Nurses’ OR ‘Physicians’ OR ‘Pharmacists’ OR ‘Allied health professionals’ OR ‘Healthcare workers’) AND (‘Review’ OR ‘Systematic review’ OR ‘Meta-analysis’ OR ‘Literature review’) AND (‘Implementation’ OR ‘Program development’)
2. (‘Internet-based education’ OR ‘E-learning’ OR ‘Telemedicine education’) AND (‘Healthcare professionals’ OR ‘Medical professionals’) AND (‘Review’ OR ‘Systematic review’ OR ‘Meta-analysis’) AND (‘Implementation’ OR ‘Program development’)

The reference lists of all the studies included were manually examined to discover additional relevant records and evaluate their eligibility.

### Study Selection

Two researchers, AT and AK, conducted the selection of studies process independently. All the duplicates were removed, and then titles and abstracts were reviewed. Full articles of reviews that are relevant to the research questions evaluated against the inclusion and exclusion criteria. Any disagreements were comprehensively discussed and resolved. The reasons for excluding studies at this stage were recorded. They will be explicitly detailed in the Preferred Reporting Items for Systematic Reviews and Meta-Analyses (PRISMA) diagram, providing transparency and clarity in the study selection process (Liberati et al., 2009).

### Quality Assessment

As the review includes qualitative studies, Enhancing Transparency in Reporting the Synthesis of Qualitative Research (ENTREQ) statement that comprises 21 items was used (Tong et al., 2012). These items are subsequently categorized into five domains, namely introduction, methods and methodology, literature search and selection, appraisal, and synthesis of findings, which helped to assess the quality of selected reviews.

### Data Extraction

A table was created to collect data from each review that was examined. This form encompassed CFIR construct, CFIR component, CFIR sub-component, sources and intervention designs. To ensure the reliability and consistency of our findings, two independent reviewers, AT and AK, conducted the data extraction process independently. Any discrepancies were addressed through discussion. Subsequently, two additional researchers, FF and VS, cross-verified the final dataset to ensure the accuracy of the extracted information.

### Synthesis of results

Themes identified from qualitative studies were directly matched with the constructs in the Consolidated Framework for Implementation Research (CFIR). CFIR was created by Damschroder et al. (2022), and it provides a structured approach for understanding and evaluating the factors influencing the successful implementation of innovations, interventions, or programs in diverse settings. The five major domains of CFIR (Intervention Characteristics, Outer Setting, Inner Setting, Characteristics of Individuals, and Process) were used to categorize and organize the extracted. Within each domain, specific constructs and sub-constructs were utilized to provide a nuanced understanding of the factors influencing the implementation of educational interventions for physicians. However, quantitative studies were transformed into themes and then were coded against the CFIR constructs. This allowed for a thorough analysis that integrated qualitative and adapted quantitative data insights. Furthermore, key themes associated with the facilitators and barriers impacting the implementation of online educational interventions were gathered also from discussion section of the papers. These sections frequently provided supplementary explanations, contributing valuable perspectives that enhanced the findings.

## Results

A total of 3340 papers were found as a result of searches as it can be seen in Figure 1. Of them, 1,862 were excluded based on the screening of the title or an abstract. Remaining 180 studies were screened as a full paper before a decision could be made. Finally, of the full papers assessed, 18 met the criteria for inclusion and were thus selected for this review.

**Figure 1.**
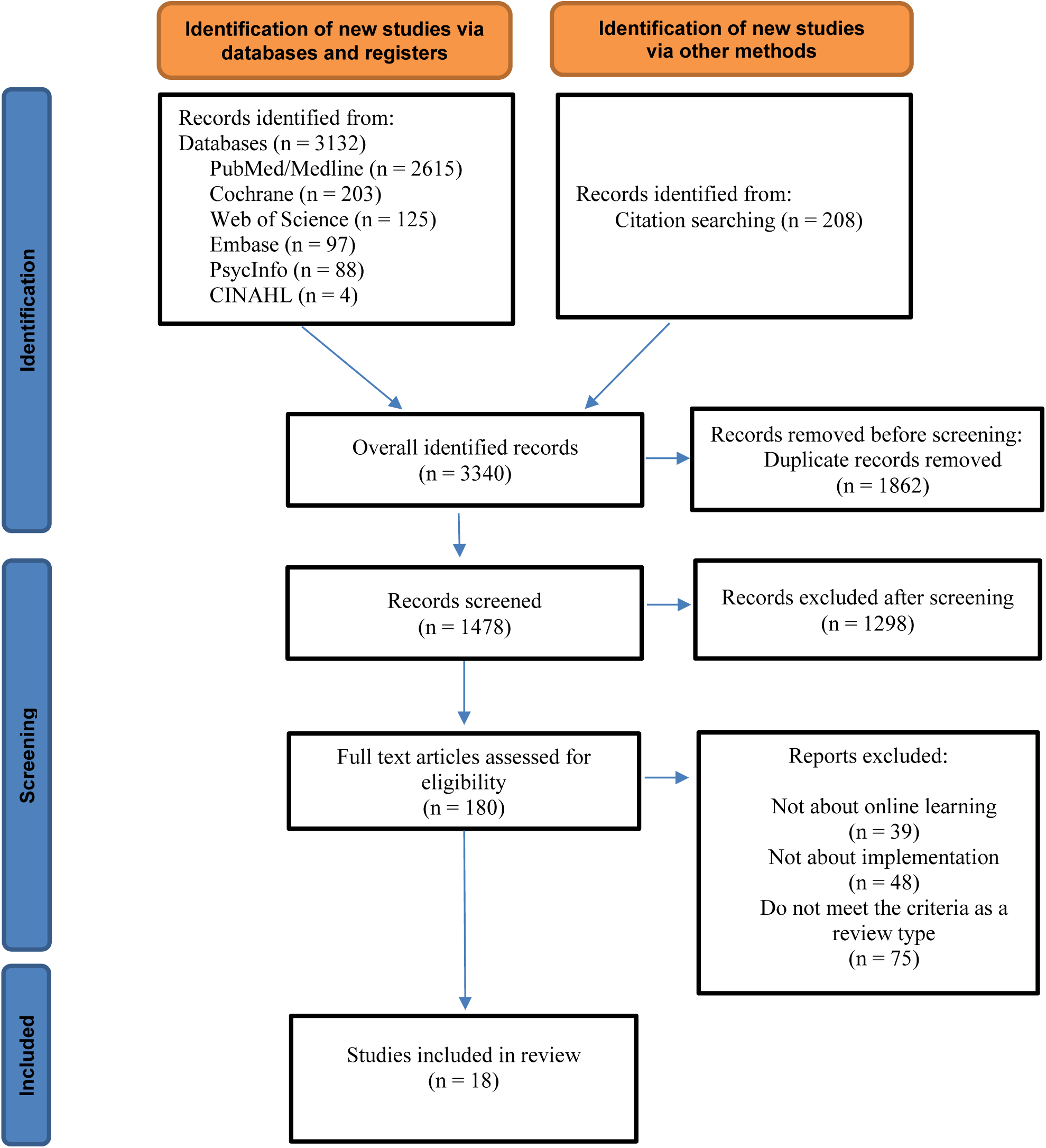
PRISMA Flow Diagram of Study Selection.

All the papers included were published between years 2018 to 2023. The settings of the studies vary and include seven medical education institutions, nine mixed settings (which involve various healthcare or educational contexts), two hospitals, one primary care, one outpatient settings, and one specific medical school named Tel Aviv Sackler Medical School. All papers were written in English (Appendix).

The synthesized information from the reviews is classified and grouped according to the main constructs and components of the CFIR framework. The parts with the most information support are explained below.

The implementation of innovations and educational interventions in medical settings is influenced by a multitude of factors.

Under the Innovation construct, facilitators such as evidence-based practices, adaptability, and innovative designs (see Table 1) are found to contribute to the successful implementation of online learning initiatives. For instance, simulation and workshops, specific skills training, and the integration of technology have been highlighted by Regmi & Jones (2020) as effective strategies to improve medical education. Similarly, Bogossian et al. (2023) emphasize the importance of addressing socialization issues and promoting cohesive approaches in interprofessional education (IPE) implementations.

**Table 1.** Implementation Barriers and Facilitators under the Innovation Construct.

| CFIR construct | CFIR component | CFIR sub-component | Sources |
| --- | --- | --- | --- |
| Innovation | Innovation Source |  |  |
|  | Innovation Evidence-Base |  | 2, 3, 10, 17 |
|  | Innovation Relative Advantage |  | 3 |
|  | Innovation Adaptability |  | 1, 5, 16 |
|  | Innovation Trialability |  |  |
|  | Innovation Complexity |  | 3, 5, 6 |
|  | Innovation Design |  | 2, 6, 7, 10, 16, 17 |
|  | Innovation Cost |  |  |

*Note.* 1. (Barde, 2020); 2. (Bogossian et al., 2023); 3. (Boutros et al., 2023); 4. (Car et al., 2022); 5. (Claflin et al., 2022); 6. (Danilovich et al., 2021); 7. (Dankner et al., 2018); 8. (Darley et al., 2022); 9. (Dawe & McKelvie, 2020); 10. (Delungahawatta et al., 2022); 11. (Eltahir et al., 2023); 12. (Hincapié et al., 2020); 13. (Joshi et al., 2021); 14. (Kho et al., 2020); 15. (Mohammadibakhsh et al., 2023); 16. (Regmi & Jones, 2020); 17. (Thomae et al., 2023); 18. (Ye et al., 2023).

Outer setting factors, including local conditions, partnerships, policies and laws (see Table 2), are stated as notably influencing the implementation process. Car et al. (2022) underscore the importance of adequate physical infrastructure and clear guidelines for digital health professions education. However, challenges such as regulatory hurdles and limited funding impede progress in this domain.

**Table 2.** Implementation Barriers and Facilitators under the Outer Setting Construct.

| CFIR construct | CFIR component | CFIR sub-component | Sources |
| --- | --- | --- | --- |
| Outer setting | Critical Incidents |  |  |
|  | Local Attitudes |  |  |
|  | Local Conditions |  | 4, 5, 6, 8, 9, 15, 17, 18 |
|  | Partnerships & Connections |  | 2, 4, 6, 9, 11, 14, 17, 18 |
|  | Policies & Laws |  | 2, 4, 5, 9, 12, 15, 18 |
|  | Financing |  | 1, 4, 18 |
|  | External Pressure |  |  |
|  |  | Societal Pressure |  |
|  |  | Market Pressure | 3 |
|  |  | Performance-Measurement Pressure |  |

*Note.* 1. (Kaur, Sethi, & Barde, 2020); 2. (Bogossian et al., 2023); 3. (Boutros et al., 2023); 4. (Car et al., 2022); 5. (Claflin et al., 2022); 6. (Danilovich et al., 2021); 7. (Dankner et al., 2018); 8. (Darley et al., 2022); 9. (Dawe & McKelvie, 2020); 10. (Delungahawatta et al., 2022); 11. (Eltahir et al., 2023); 12. (Hincapié et al., 2020); 13. (Joshi et al., 2021); 14. (Kho et al., 2020); 15. (Mohammadibakhsh et al., 2023); 16. (Regmi & Jones, 2020); 17. (Thomae et al., 2023); 18. (Ye et al., 2023).

Within the Inner setting, structural characteristics such as physical and information technology infrastructure, communication, and cultural aspects, particularly recipient-centeredness and learning-centeredness, are identified as crucial factors in implementing online educational interventions for physicians (see Table 3). Additionally, tension for change and resource availability, including finance, time, and materials, are highlighted as key factors influencing implementation success. For instance, Claflin et al. (2022) emphasize the importance of adaptability and ongoing support for facilitators in health education interventions, while Danilovich et al. (2021) underscore the need for a comprehensive assessment of online education systems in family medicine residency programs. This underscores the significance of addressing structural, cultural, and resource-related considerations in effectively implementing online educational interventions within medical settings.

**Table 3.** Implementation Barriers and Facilitators under the Inner Setting Construct.

| CFIR construct | CFIR component | CFIR sub-component | Sources |
| --- | --- | --- | --- |
| Inner setting | Structural Characteristics |  |  |
|  |  | Physical Infrastructure | 1, 3, 4, 7, 8, 11, 17, 18 |
|  |  | Information Technology Infrastructure | 1, 3, 8, 10, 11, 12, 14, 16, 17, 18 |
|  |  | Work Infrastructure | 6, 7, 8, 13, 14, 15 |
|  | Relational Connections |  |  |
|  | Communications |  | 1, 4, 9, 11, 13, 14 |
|  | Culture |  | 1 |
|  |  | Human Equality-Centeredness | 18 |
|  |  | Recipient-Centeredness | 2, 4, 6, 7, 8, 9, 12, 13, 14, 15, 16 |
|  |  | Deliverer-Centeredness |  |
|  |  | Learning-Centeredness | 6, 9, 16 |
|  | Tension for Change |  | 4, 7, 9, 10, 13, 14 |
|  | Compatibility |  |  |
|  | Relative Priority |  |  |
|  | Incentive Systems |  |  |
|  | Mission Alignment |  |  |
|  | Available Resources |  |  |
|  |  | Funding | 2, 4, 5, 6, 7, 8, 9, 10, 12, 13, 14, 15, 16, 17 |
|  |  | Space | 4, 5, 6, 7, 8, 9, 14, 16, 17 |
|  |  | Materials & Equipment | 5, 8 |
|  | Access to Knowledge & Information |  | 4, 5 |

*Note.* 1. (Kaur, Sethi, & Barde, 2020); 2. (Bogossian et al., 2023); 3. (Boutros et al., 2023); 4. (Car et al., 2022); 5. (Claflin et al., 2022); 6. (Danilovich et al., 2021); 7. (Dankner et al., 2018); 8. (Darley et al., 2022); 9. (Dawe & McKelvie, 2020); 10. (Delungahawatta et al., 2022); 11. (Eltahir et al., 2023); 12. (Hincapié et al., 2020); 13. (Joshi et al., 2021); 14. (Kho et al., 2020); 15. (Mohammadibakhsh et al., 2023); 16. (Regmi & Jones, 2020); 17. (Thomae et al., 2023); 18. (Ye et al., 2023).

Individual-level factors, including leadership engagement, motivation, and capability (see Table 4), are identified to shape the implementation of e-learning. Joshi et al. (2021) emphasize the importance of leadership engagement and sufficient resources in online medical education, while Delungahawatta et al. (2022) discuss the significance of interactive and asynchronous e-learning interventions, alongside challenges like financial barriers and resistance to change.

**Table 4.** Implementation Barriers and Facilitators under the Individuals Construct.

| CFIR construct | CFIR component | CFIR sub-component | Sources |
| --- | --- | --- | --- |
| Individuals | High-Level Leaders |  |  |
|  | Mid-level Leaders |  |  |
|  | Opinion Leaders |  | 12 |
|  | Implementation Facilitators |  |  |
|  | Implementation Leads |  | 4, 5, 9, 13, 14, 15, 17 |
|  | Implementation Team Members |  |  |
|  | Other Implementation Support |  | 19 |
|  | Innovation Deliverers |  |  |
|  | Innovation Recipients |  |  |
|  | Characteristics | Need |  |
|  |  | Capability | 1, 4, 5, 6, 9, 10, 14, 15, 17 |
|  |  | Opportunity |  |
|  |  | Motivation | 9, 10, 11, 16, 17 |

*Note.* 1. (Kaur, Sethi, & Barde, 2020); 2. (Bogossian et al., 2023); 3. (Boutros et al., 2023); 4. (Car et al., 2022); 5. (Claflin et al., 2022); 6. (Danilovich et al., 2021); 7. (Dankner et al., 2018); 8. (Darley et al., 2022); 9. (Dawe & McKelvie, 2020); 10. (Delungahawatta et al., 2022); 11. (Eltahir et al., 2023); 12. (Hincapié et al., 2020); 13. (Joshi et al., 2021); 14. (Kho et al., 2020); 15. (Mohammadibakhsh et al., 2023); 16. (Regmi & Jones, 2020); 17. (Thomae et al., 2023); 18. (Ye et al., 2023).

In the Implementation process domain, key components such as engaging, reflecting and evaluating, and adapting are identified as important factors in implementation (see Table 5). For example, Kaur, Sethi, & Barde (2020) emphasize the significance of engaging participants and soliciting their feedback on the intervention, while Claflin et al. (2022) highlight the adaptability of developed e-learning initiatives. This underscores the importance of active involvement, continuous assessment, and flexibility in the implementation process to ensure the effectiveness and sustainability of online educational interventions for physicians.

**Table 5.** Implementation Barriers and Facilitators under the Implementation Process Domain Construct.

| CFIR construct | CFIR component | CFIR sub-component | Sources |
| --- | --- | --- | --- |
| Implementation Process Domain | Teaming |  |  |
|  | Assessing Needs | Innovation Deliverers | 11 |
|  |  | Innovation Recipients |  |
|  | Assessing Context |  |  |
|  | Planning |  | 17 |
|  | Tailoring Strategies |  | 17 |
|  | Engaging | Innovation Deliverers |  |
|  |  | Innovation Recipients | 1, 3, 11, 16 |
|  | Doing |  |  |
|  | Reflecting & Evaluating | Implementation | 2, 5, 6, 7, 10 |
|  |  | Innovation |  |
|  | Adapting |  | 5, 16 |

*Note.* 1. (Kaur, Sethi, & Barde, 2020); 2. (Bogossian et al., 2023); 3. (Boutros et al., 2023); 4. (Car et al., 2022); 5. (Claflin et al., 2022); 6. (Danilovich et al., 2021); 7. (Dankner et al., 2018); 8. (Darley et al., 2022); 9. (Dawe & McKelvie, 2020); 10. (Delungahawatta et al., 2022); 11. (Eltahir et al., 2023); 12. (Hincapié et al., 2020); 13. (Joshi et al., 2021); 14. (Kho et al., 2020); 15. (Mohammadibakhsh et al., 2023); 16. (Regmi & Jones, 2020); 17. (Thomae et al., 2023); 18. (Ye et al., 2023).

## Discussion

The conversation about incorporating online learning into medical education reveals a complex mix of factors that impact its successful implementation. To effectively integrate online learning, it is crucial to embrace innovations that are evidence-based and adaptable. However, various external challenges, such as technological and limited funding, must be navigated to ensure smooth adoption. Within organizations, creating a supportive culture and establishing comprehensive assessment systems are vital components of success. Yet, ongoing challenges include addressing resistance to change, tailoring interventions to community needs, and ensuring adequate resources. Looking ahead, collaborative efforts involving stakeholders from different levels are indispensable to foster an environment that supports innovation and drives significant improvements in medical education through online learning.

A key challenge identified by Adedoyın & Soykan (2020) is limited access to reliable internet connectivity and outdated hardware, which can significantly impede the effectiveness of online learning experiences, especially in low- and middle-income countries. This aligns with our research findings that highlight the importance of robust technological infrastructure for online learning adoption. Ensuring that institutions have the necessary technology resources, from reliable internet to modern hardware, is essential for the success of online education initiatives. Institutional resistance and lack of support represent another barrier to the effective implementation of online learning. Bury et al. (2006) found that resistance to change within institutions can significantly hinder the adoption of online interventions. This observation resonates with the broader research results, which emphasize the need for a supportive organizational culture that embraces innovation. Addressing resistance to change and fostering a culture that supports innovation are critical steps in overcoming institutional barriers to online learning.

The results of the study suggest that customized implementation techniques that make use of evidence-based practices, flexibility, and creative designs are necessary to improve the performance of online learning programs in healthcare environments. Fostering an environment that is favorable to implementation success requires addressing both inner and outside setting variables, such as physical and cultural characteristics, as well as local conditions, collaborations, and policies. Implementation efforts can also be strengthened by placing a higher priority on individual-level elements like motivation and leadership engagement, as well as by using efficient implementation procedures that highlight stakeholder interaction and flexibility. In order to determine how well these suggestions will work to improve doctors’ adoption and sustainability of online learning programs, more investigation and assessment are necessary in the future. This will help to advance both medical education and healthcare delivery.

The study has a number of advantages and disadvantages. The utilization of pre-existing evaluations poses a possible constraint as the quality and technique may differ, thereby impacting the dependability of the combined results. Publication bias may have an impact on the outcomes of the studies that are included in systematic reviews. Furthermore, even though the CFIR framework is useful for examining implementation issues, it could miss some subtleties and complexities that are specific to each setting. However the study also provides several advantages. Conducting a methodical literature study can enhance the reliability and repeatability of results. A thorough method of examining obstacles and enablers to the implementation of online educational interventions for physicians is to employ the CFIR framework. Overall, despite its limitations, the study offers valuable insights that can inform future research and practice in medical education.

Future investigations should concentrate on longitudinal studies to monitor the long-term efficacy of online medical education programs. It is essential to comprehend how technology may support successful online learning, investigate cultural aspects that influence implementation, and create specialized implementation techniques. Additionally, to produce high-quality evidence for well-informed decision-making in medical education, rigorous evaluation techniques like randomized controlled trials are required. We also suggest policy recommendations that include funding for evaluation studies, prioritizing investments in digital infrastructure, encouraging interprofessional education, enforcing clear regulatory frameworks, and establishing supportive policies for digital health education. These regulations seek to improve patient outcomes and healthcare delivery by guaranteeing the efficacy, accessibility, and quality of online medical education.

## Data Availability

All data produced in the present study are available upon reasonable request to the authors

---

**Supplement 1.**
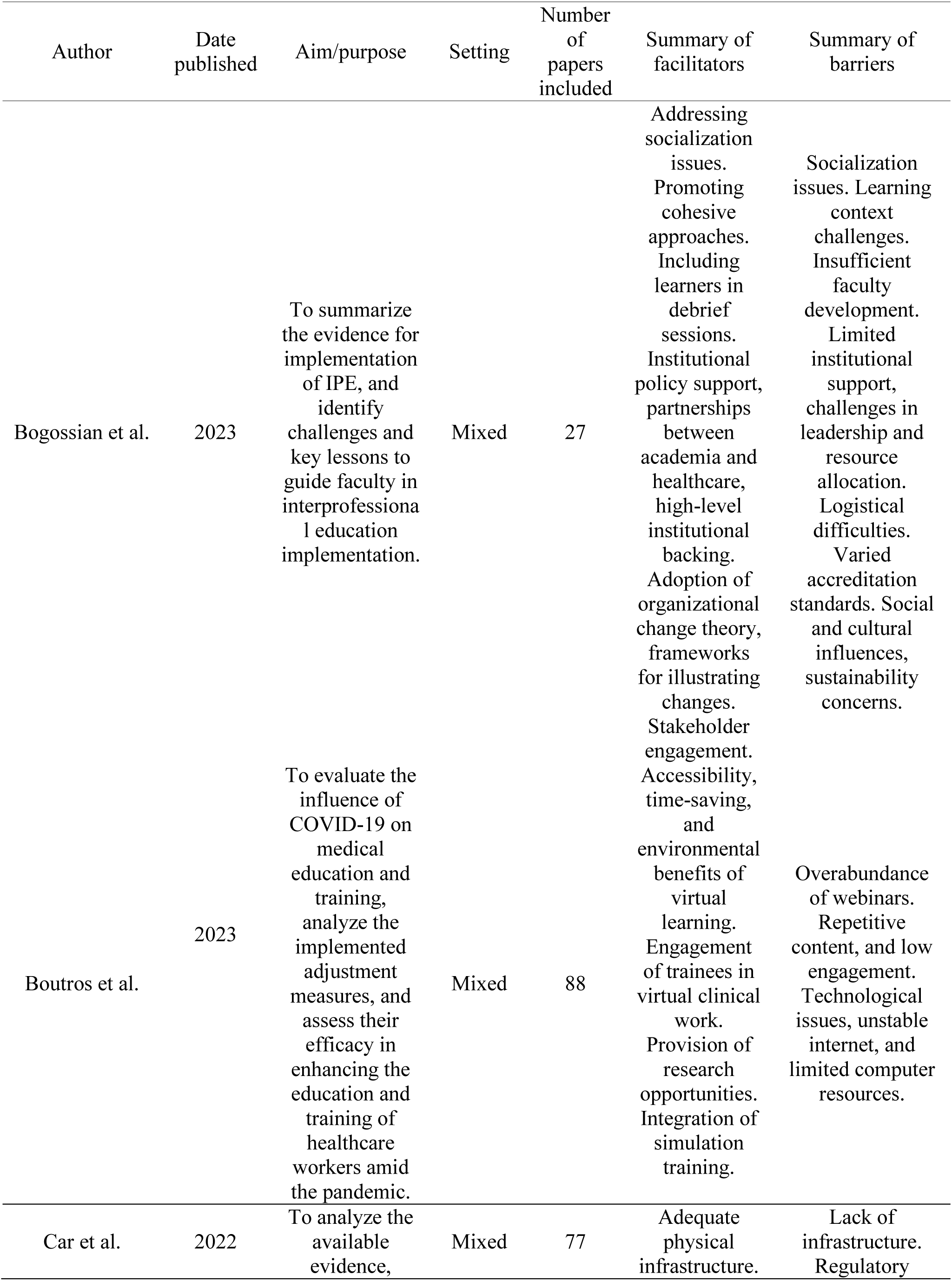
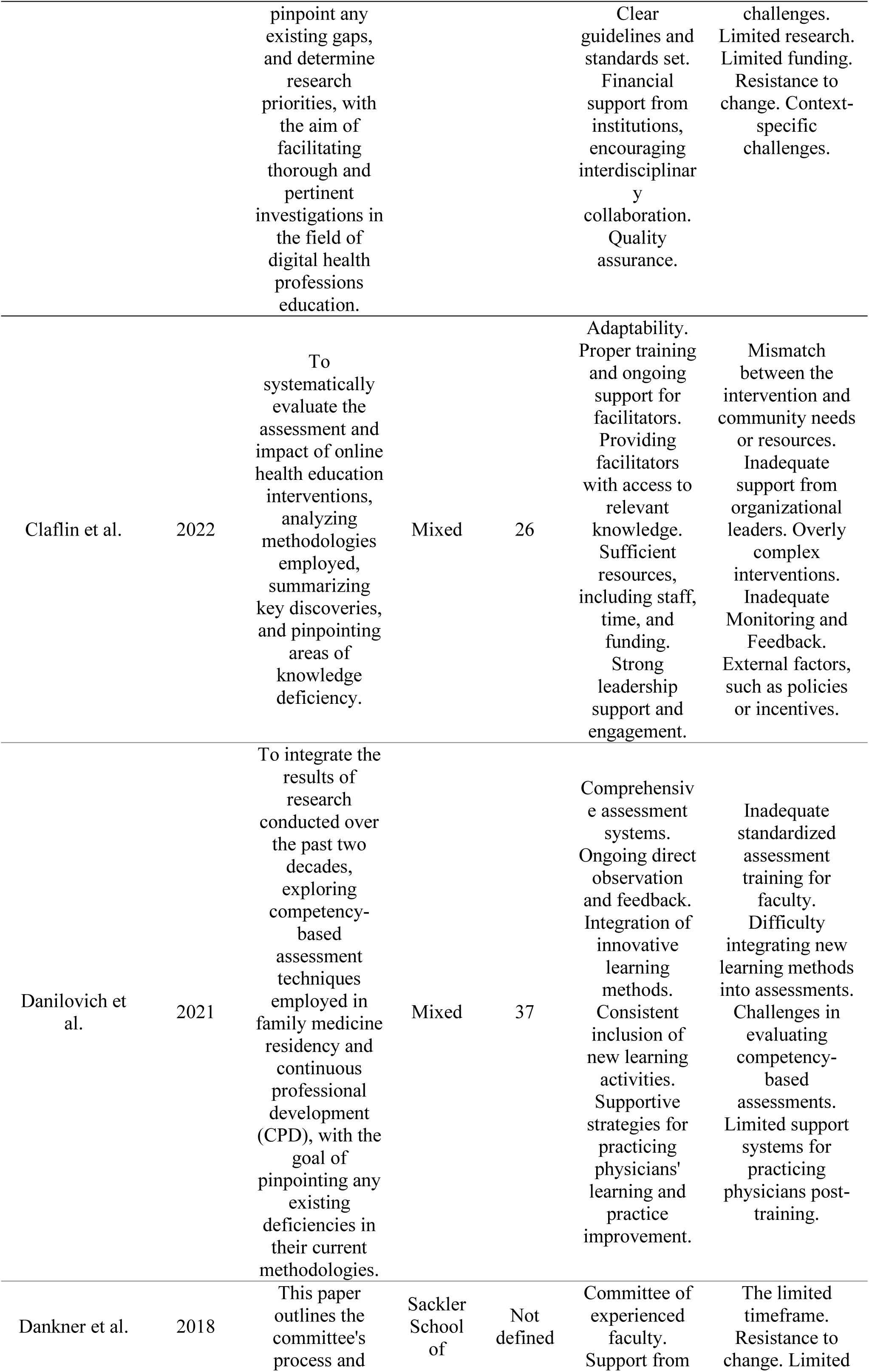
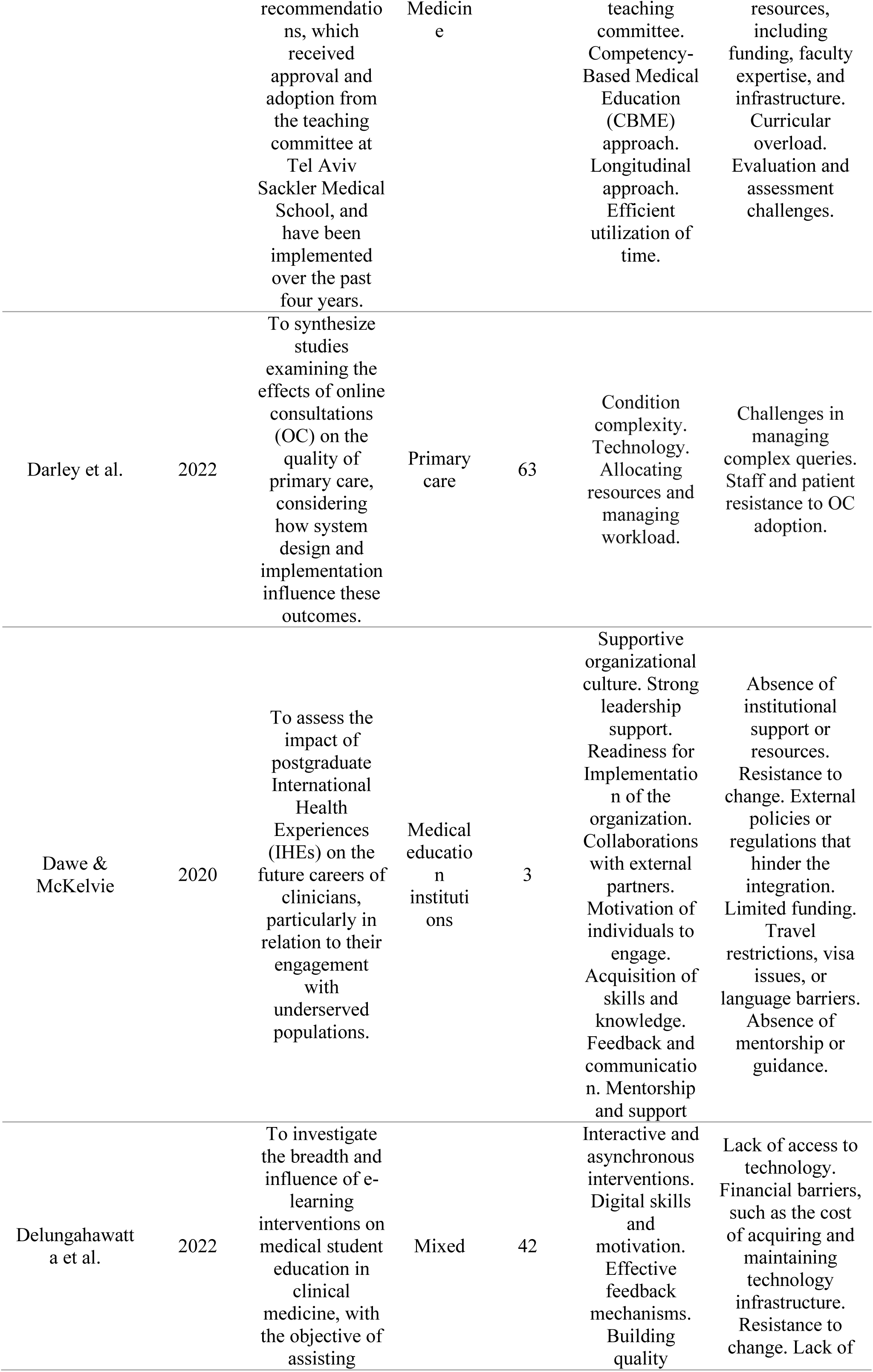
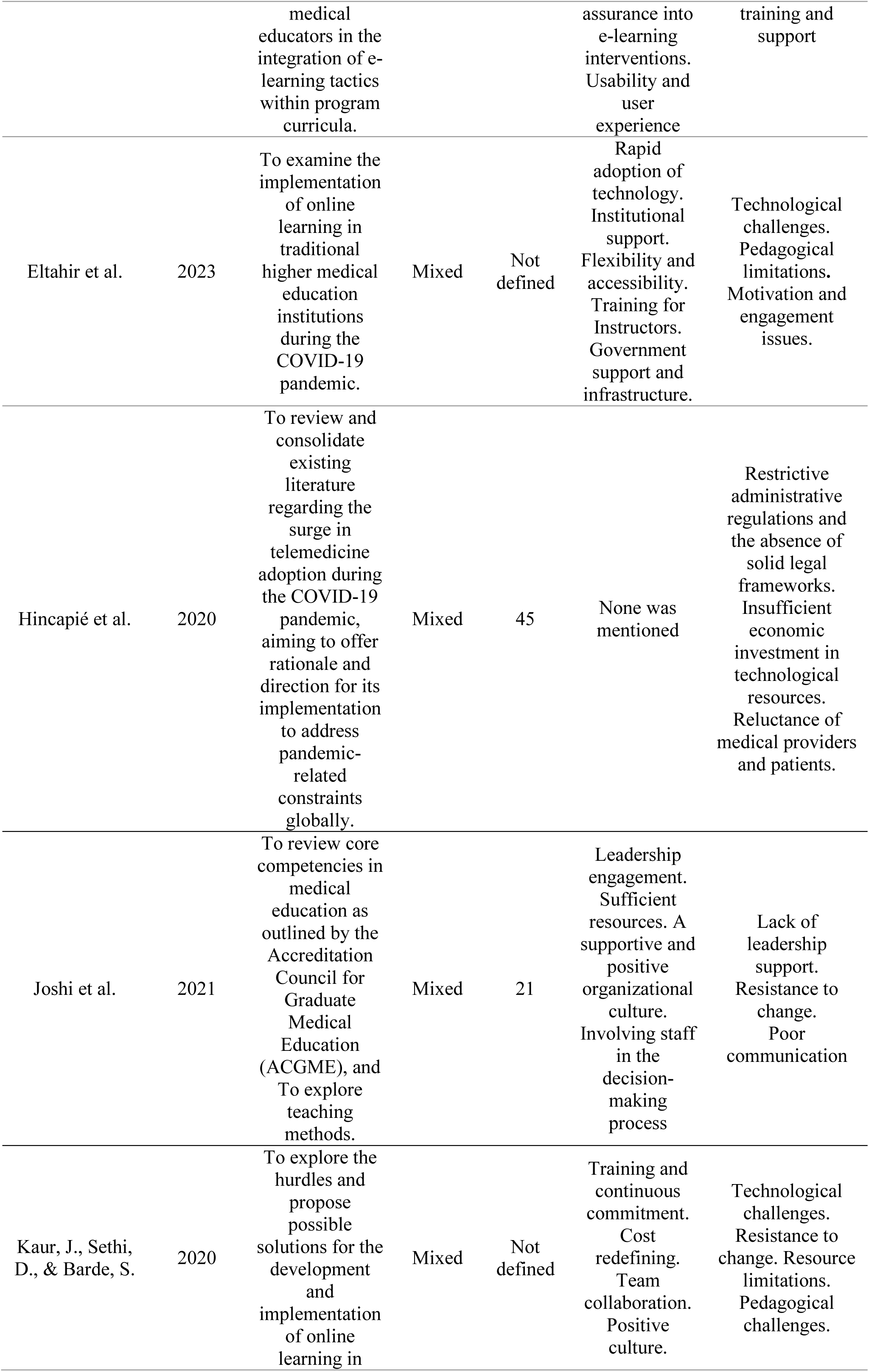
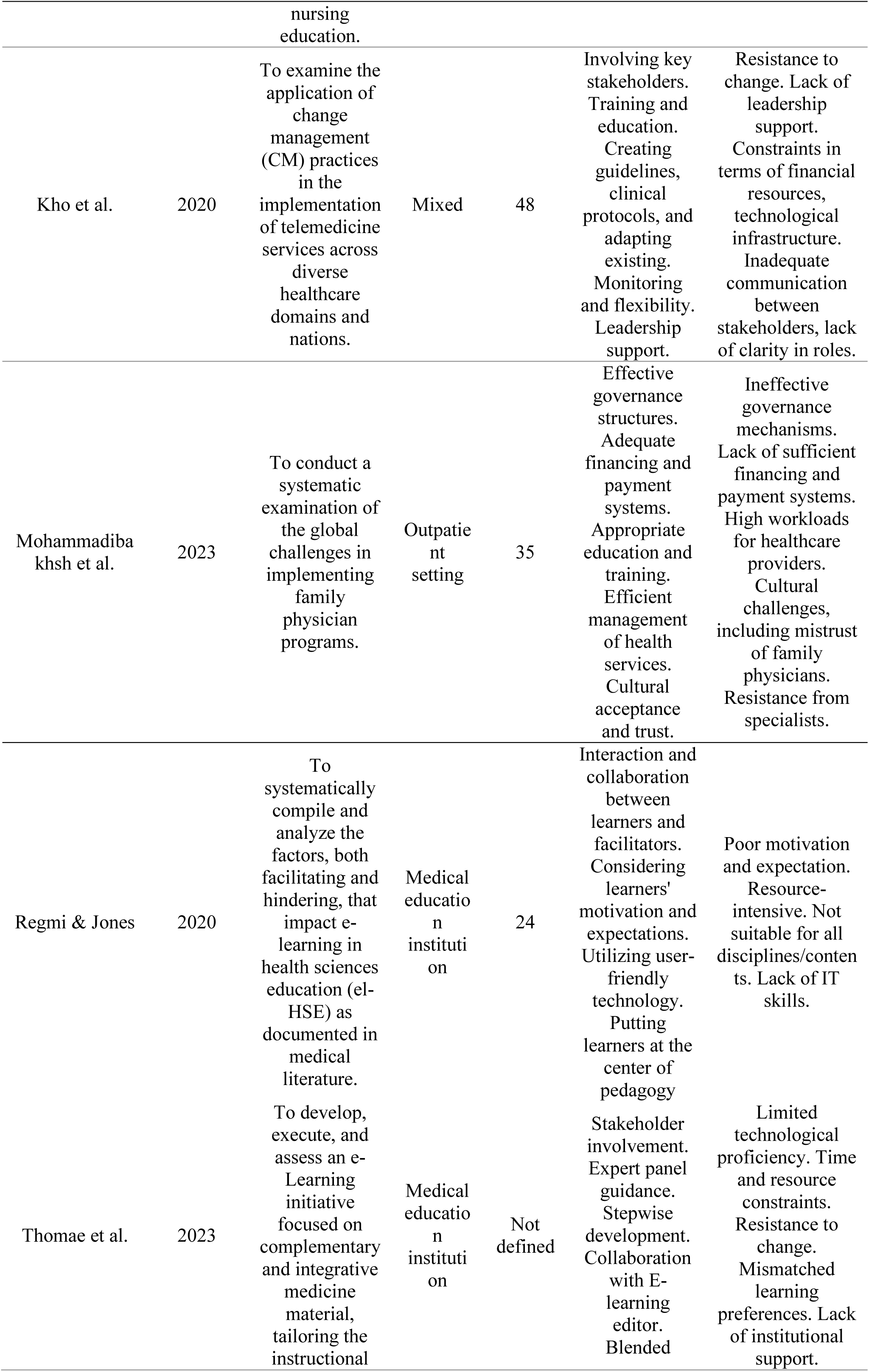
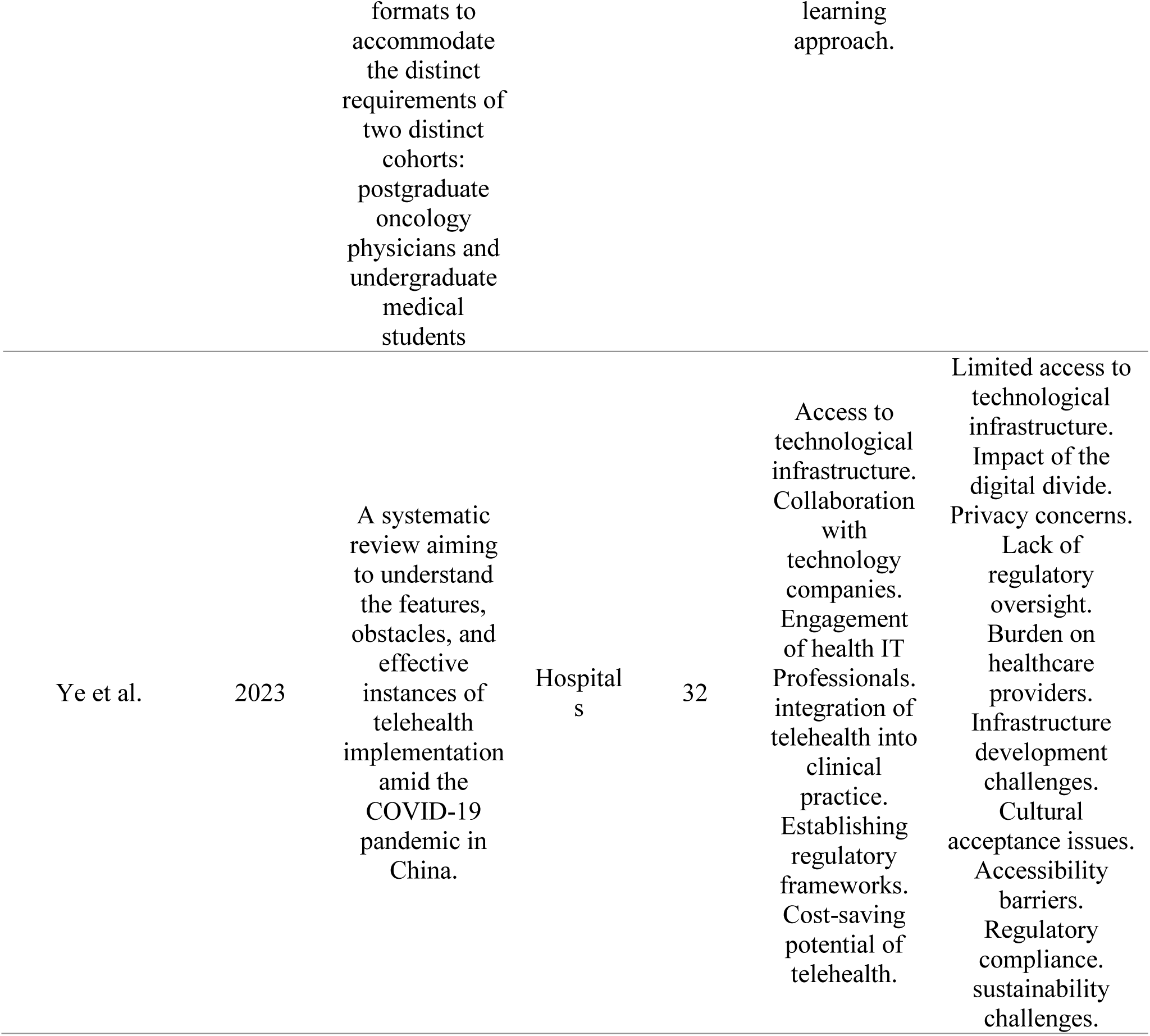
Summary Details of the 18 Included Studies.

